# Phylodynamic analysis reveals disparate transmission dynamics of *Mycobacterium tuberculosis*-complex lineages in Botswana

**DOI:** 10.1101/2024.10.31.24316225

**Authors:** Qiao Wang, Ivan Barilar, Volodymyr M. Minin, Chawangwa Modongo, Patrick K. Moonan, Alyssa Finlay, Rosanna Boyd, John E. Oeltmann, Tuduetso L. Molefi, Nicola M. Zetola, Timothy F. Brewer, Stefan Niemann, Sanghyuk S. Shin

**Author notes:** These authors have contributed equally to this work (Stefan Niemann, Sanghyuk S. Shin).

## Abstract

Tuberculosis epidemics have traditionally been conceptualized as arising from a single uniform pathogen. However, *Mycobacterium tuberculosis*-complex (Mtbc), the pathogen causing tuberculosis in humans, encompasses multiple lineages exhibiting genetic and phenotypic diversity that may be responsible for heterogeneity in TB transmission. We analysed a population-based dataset of 1,354 Mtbc whole-genome sequences collected over four years in Botswana, a country with high HIV and tuberculosis burden. We identified Lineage 4 (L4) as the most prevalent (87.4%), followed by L1 (6.4%), L2 (5.3%), and L3 (0.9%). Within L4, multiple sublineages were identified, with L4.3.4 being the predominant sublineage. Phylodynamic analysis revealed L4.3.4 expanded steadily from late 1800s to early 2000s. Conversely, L1, L4.4, and L4.3.2 showed population trajectories closely aligned with the HIV epidemic. Meanwhile, L2 saw rapid expansion throughout most of the 20^th^ century but declined sharply in early 1990s. Additionally, pairwise genome comparison of Mtbc highlighted differences in clustering proportions due to recent transmission at the sublineage level. These findings emphasize the diverse transmission dynamics of strains of different Mtbc lineages and highlight the potential for phylodynamic analysis of routine sequences to refine our understanding of lineage-specific behaviors.

## Introduction

Despite being preventable and curable, tuberculosis (TB) claims an estimated 1.4 million lives each year worldwide, surpassing deaths caused by any other infectious disease besides severe acute respiratory syndrome coronavirus-2 (SARS-CoV-2).^1^ The SARS-CoV-2 pandemic highlighted the public health importance of monitoring the spread of different genomic variants globally and in local settings.^2^ Indeed, genomic epidemiology has been instrumental in outbreak investigation and public health over the previous decade by tracing the spread of various infectious pathogens and informing key stakeholders. For instance, genomic surveillance of viral pathogens guided Ebola vaccine allocation in west Africa during the 2013–2016 outbreak,^3^ identified likely source and routes of zika virus introductions into the United States during the 2016 outbreak,^4^ and has continuously mapped transmission networks and detected high-risk groups in the ongoing HIV pandemic.^5, 6^ These findings are facilitated through the use of phylodynamics, a statistical framework that integrates high-resolution genetic data with individual-level epidemiological data to shed light on the transmission dynamics of infectious pathogens, providing timely guidance for public health interventions.^7, 8^

For TB research, large-scale comparative genomic analysis have provided valuable insights into the origin and genetic diversity of the human-adapted TB bacteria, collectively known as the *Mycobacterium tuberculosis*–complex (Mtbc).^9, 10^ There are currently a total of 9 major lineages of Mtbc and 64 sublineages that characterize their genetic diversity according to a 62-single nucleotide polymorphism (SNP) barcode.^11, 12, 13^ Strain diversity in Mtbc can lead to important phenotypic variations in virulence, antibiotic resistance patterns, and transmissibility.^14,15^ For example, Lineages 2 and 4 are widespread globally and noted for their enhanced transmissibility, as evidenced by comparing Mtbc genomes (*e.g.,* genotypic clustering proportions) collected in many settings.^16, 17^ By contrast, Lineage 1 is less likely to cause disease shortly after infection and may have a reduced ability to spread.^14, 16, 17^ However, most epidemiological studies and surveillance efforts still do not distinguish between Mtbc strains, and the conventional approach of treating TB as a uniform epidemic fails to account for pathogen heterogeneity and potential differences in the transmission dynamics of Mtbc strains, which could vary significantly across different regions and populations.^14, 18, 19^ Much like the insights for viral transmission dynamics gained from genomic surveillance of SARS-CoV-2 variants,^2^ analysing Mtbc sequences to uncover long-term epidemiological trends of specific Mtbc strains may offer a more granular understanding of the TB transmission dynamics at the population level, which could improve our ability to predict the trajectory of the TB epidemic and design more tailored interventions for a given setting.

Located in southern Africa, Botswana continues to face one of the most severe TB endemics globally and remains one of the 30 high TB/HIV burden countries.^1^ Despite the public health implications, the evolutionary history and transmission dynamics of Mtbc lineages remain poorly understood in this high burden country. In this study, we sought to expand our understanding of the Mtbc population dynamics underlying the long-standing TB endemic in Botswana by leveraging a countrywide dataset of whole genome sequences (WGS) from Mtbc isolates collected over four years.^20^ Our aim was to delineate the long-term spread of Mtbc subpopulations in the context of sociohistorical events and the generalized HIV epidemic in the region. In addition to the broader long-term trends revealed through phylodynamic analysis, we also examined clustering of cases by Mtbc lineages based on pairwise genetic distances to track recent transmission patterns. We hypothesized that TB in Botswana is composed of a collection of heterogeneous epidemics driven by distinct transmission dynamics of Mtbc lineages.

## Results

### Distribution of Mtbc lineages in Botswana

During the study period, 1,426 Mtbc isolates underwent successful WGS and were assigned into known lineages based on lineage-specific SNPs. We excluded 72 isolates with evidence of mixed Mtbc strain infection for subsequent phylogenetic and phylodynamic analysis. Among the remaining 1,354 isolates, 1,184 belonged to lineage 4 (L4; 87.4%), 86 (6.4%) were classified as lineage 1 (L1), 72 (5.3%) were lineage 2 (L2), and 12 (0.9%) were lineage 3 (L3). L4 was found to encompass a number of sublineages, including L4.1 (n=2; 0.1%), L4.1.1 (n=141; 10.4%), L4.1.2 (n=149; 11.0%), L4.2.2 (n=1; 0.1%), L4.3.2 (n=117; 8.6%), L4.3.3 (n=27; 2.0%), L4.3.4 (n=400; 29.5%), L4.4 (n=189; 14.0%), L4.6 (n=2; 0.1%), L4.7 (n=14; 1.0%), L4.8 (n=92; 6.8%), and L4.9 (n=50; 3.7%). Within L1, 22 (1.6%) were L1.1 and 64 (4.7%) were L1.2. All of the L2 isolates were classified as L2.2 (Figure 1). Furthermore, Gaborone harbored more Mtbc strain diversity compared to Ghanzi. L4.3.4, the most prevalent sublineage, was found in higher proportion in Ghanzi than Gaborone (67% vs. 21%; Table 1). Host characteristics were similar among the major Mtbc lineages (Table 2).

**Figure 1.**
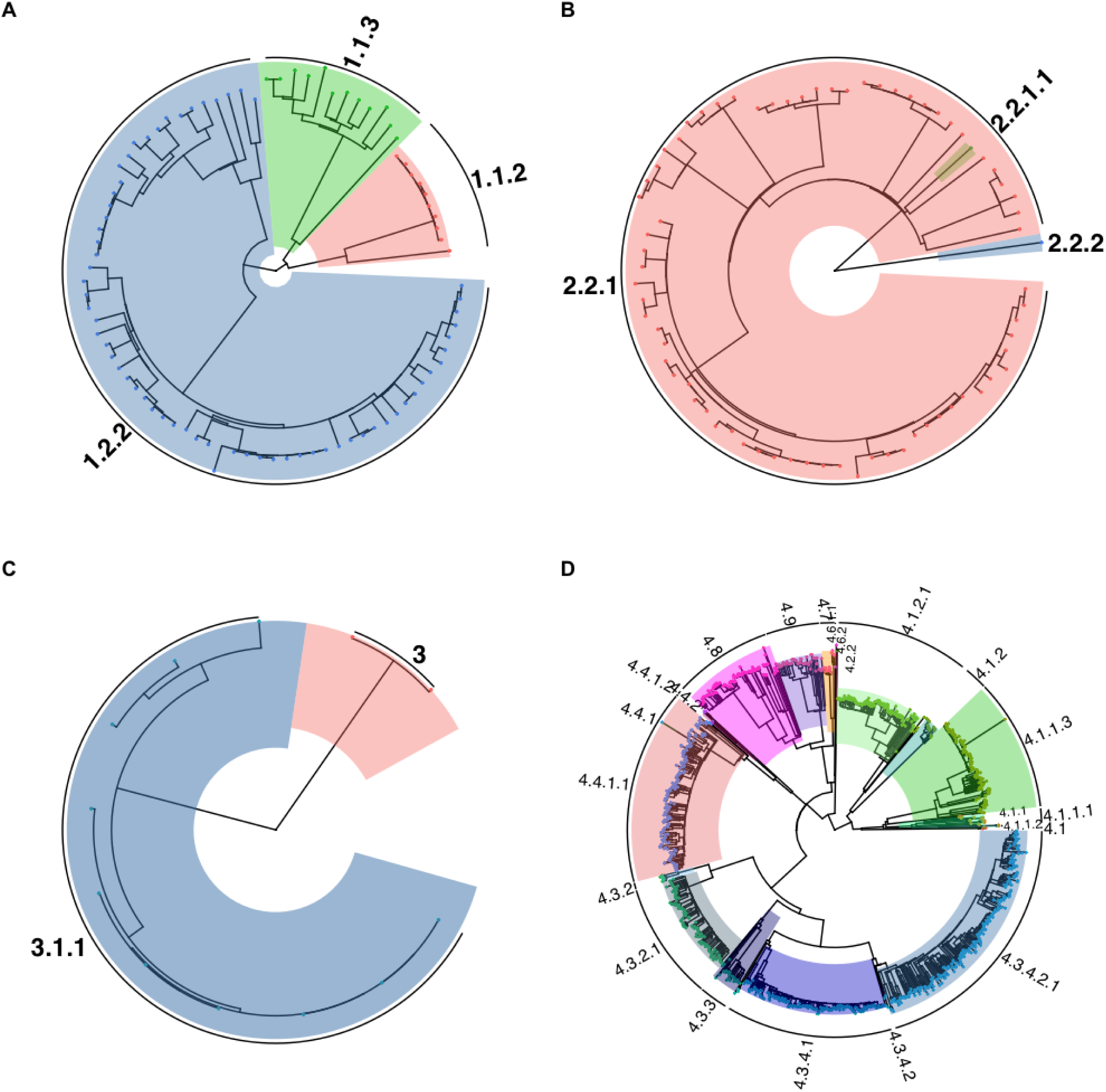
Maximum likelihood phylogeny of *Mycobacterium tuberculosis*–complex (Mtbc) isolates collected in Botswana, 2012–2016 (n = 1,354). A. Phylogeny of Lineage 1 Indo-Oceanic Mtbc isolates (n = 86). B. Phylogeny of Lineage 2 East Asian Mtbc isolates (n = 72). C. Phylogeny of Lineage 3 East African-Indian Mtbc isolates (n = 12). D. Phylogeny of Lineage 4 Euro-American Mtbc isolates (n = 1184)

**Table 1.** Distribution of *Mycobacterium tuberculosis* complex lineages by district in Botswana, 2012–2016.

|  | District |  | Total<br>n (%) |
| --- | --- | --- | --- |
|  | Gaborone<br>n (%) | Ghanzi<br>n (%) |  |
| **Lineage 1 Indo-Oceanic** | 83 (7.6) | 3 (1.2) | 86 (6.4) |
| L1.1 | 22 (2.0) | 0 | 22 (1.6) |
| L1.2 | 61 (5.6) | 3 (1.2) | 64 (4.7) |
| **Lineage 2 East Asian** |  |  |  |
| L2.2 | 65 (5.9) | 7 (2.7) | 72 (5.3) |
| **Lineage 3 East African-Indian** | 12 (1.1) | 0 | 12 (0.9) |
| **Lineage 4 Euro-American** | 938 (85.4) | 246 (96.1) | 1184 (87.4) |
| L4.1 | 2 (0.2) | 0 | 2 (0.1) |
| L4.1.1 | 136 (12.4) | 5 (2.0) | 141 (10.4) |
| L4.1.2 | 129 (11.7) | 20 (7.8) | 149 (11.0) |
| L4.2.2 | 1 (0.1) | 0 | 1 (0.1) |
| L4.3.2 | 107 (9.7) | 10 (3.9) | 117 (8.6) |
| L4.3.3 | 24 (2.2) | 3 (1.2) | 27 (2.0) |
| L4.3.4 | 229 (20.9) | 171 (66.8) | 400 (29.5) |
| L4.4 | 164 (14.9) | 25 (9.8) | 189 (14.0) |
| L4.6 | 2 (0.2) | 0 | 2 (0.1) |
| L4.7 | 13 (1.2) | 1 (0.4) | 14 (1.0) |
| L4.8 | 84 (7.7) | 8 (3.1) | 92 (6.8) |
| L4.9 | 47 (4.3) | 3 (1.2) | 50 (3.7) |
| **Total** | 1098 (100) | 256 (100) | 1354 (100) |

### Historical origins and population dynamics of Mtbc lineages

Demographic reconstruction was performed for L1, L2, L4.1.1, L4.1.2, L4.3.2, L4.3.4, L4.4, and L4.8. The time to the most recent common ancestor (MRCA) refers to the estimated age of a common ancestral population from which current populations or strains of Mtbc have descended. We inferred the MRCA to have emerged around year 1727 (95% highest posterior density interval [HPDI], 1607–1828) and 1900 (95% HPDI 1854–1938) for strains of L1 and L2, respectively. Time to MRCA varied among L4 sublineages, ranging from year 1695 for L4.1.2 (95% HPDI 1546–1817) to 1941 for L4.3.2 (95% HPDI 1911–1966; Table 3). Our analysis revealed that strains of L4.3.4 experienced a steady expansion from late 1800s to early 2000s, and remained the most prevalent sublineage in recent years. Similar population trajectory was observed for L1 and L4.4 strains, with a substantial expansion observed from the late 1970s for two decades, followed by a continuous decline from the late 1990s. For L4.3.2 strains, we observed significant growth between 1980 to early 2000s, followed by a sharp reduction afterward. L2 strains experienced a steady increase in population growth between 1940 and early 1990s, followed by a steep decline until 2000, after which the population plateaued followed by a second decline around 2012. Other L4 sublineages experienced several waves of expansion and contraction. Except for L4.1.1, all other Mtbc lineages underwent contraction after early 2000s (Figure 2 and Supplementary Figure 1). For the inferred clock rate and maximum clade credibility trees of these lineages, refer to Supplementary Table 1 and Supplementary Figure 2a-h. Genomic clustering analysis

**Table 2.** Participant characteristics by *Mycobacterium tuberculosis*–complex (Mtbc) lineages in Botswana, 2012–2016.

|  | Mtb lineage |  |  |  | Total<br>n (%) |
| --- | --- | --- | --- | --- | --- |
|  | L1<br>n (%) | L2<br>n (%) | L3<br>n (%) | L4<br>n (%) |  |
| **Sex** |  |  |  |  |  |
| Female | 35 (40.7) | 38 (52.8) | 6 (50.0) | 522 (44.1) | 601 (44.4) |
| Male | 51 (59.3) | 34 (47.2) | 6 (50.0) | 662 (55.9) | 753 (55.6) |
| **Age** |  |  |  |  |  |
| ≤15 | 2 (2.3) | 3 (4.2) | 1 (8.3) | 21 (1.8) | 27 (2.0) |
| 16-24 | 10 (11.6) | 9 (12.5) | 1 (8.3) | 226 (19.1) | 246 (18.2) |
| 25-40 | 47 (54.7) | 46 (63.9) | 7 (58.3) | 632 (53.4) | 732 (54.1) |
| 41-64 | 24 (27.9) | 14 (19.4) | 3 (25.0) | 280 (23.6) | 321 (23.7) |
| ≥65 | 3 (3.5) | 0 | 0 | 25 (2.1) | 28 (2.1) |
| **Previous TB** |  |  |  |  |  |
| No | 63 (73.3) | 57 (79.2) | 10 (83.3) | 967 (81.7) | 1097 (81.0) |
| Yes | 23 (26.7) | 15 (20.8) | 2 (16.7) | 217 (18.3) | 257 (19.0) |
| **Isoniazid preventive therapy** |  |  |  |  |  |
| No | 83 (96.5) | 64 (88.9) | 12 (100.0) | 1127 (95.2) | 1286 (95.0) |
| Yes | 2 (2.3) | 5 (6.9) | 0 | 42 (3.5) | 49 (3.6) |
| Unknown | 1 (1.2) | 3 (4.2) | 0 | 15 (1.3) | 19 (1.4) |
| **AFB smear results** |  |  |  |  |  |
| Negative | 5 (5.8) | 7 (9.7) | 3 (25.0) | 76 (6.4) | 91 (6.7) |
| Positive | 61 (70.9) | 46 (63.9) | 8 (66.7) | 752 (63.5) | 867 (64.0) |
| Not done | 20 (23.3) | 19 (26.4) | 1 (8.3) | 356 (30.1) | 396 (29.2) |
| **TB type** |  |  |  |  |  |
| Pulmonary | 79 (91.9) | 65 (90.3) | 11 (91.7) | 1075 (90.8) | 1230 (90.8) |
| Extrapulmonary | 7 (8.1) | 6 (8.3) | 0 | 105 (8.9) | 118 (8.7) |
| Unknown | 0 | 1 (1.4) | 1 (8.3) | 4 (0.3) | 6 (0.4) |
| **HIV status** |  |  |  |  |  |
| Negative | 27 (31.4) | 27 (37.5) | 5 (41.7) | 533 (45.0) | 592 (43.7) |
| Positive | 56 (65.1) | 42 (58.3) | 7 (58.3) | 616 (52.0) | 721 (53.2) |
| Unknown | 3 (3.5) | 3 (4.2) | 0 | 35 (3.0) | 41 (3.0) |
| **ART status<sup>1</sup>** |  |  |  |  |  |
| Never taken ART | 26 (46.4) | 17 (40.5) | 1 (14.3) | 278 (45.1) | 322 (44.7) |
| Taking ART | 25 (44.6) | 20 (47.6) | 3 (42.9) | 277 (45.0) | 325 (45.1) |
| Took ART but stopped | 1 (1.8) | 1 (2.4) | 2 (28.6) | 11 (1.8) | 15 (2.1) |
| Unknown | 4 (7.1) | 4 (9.5) | 1 (14.3) | 50 (8.1) | 59 (8.2) |

Abbreviations: AFB: acid-fast bacilli. ART: antiretroviral therapy.
<sup>1</sup>Among HIV-positive individuals only.

**Table 3.** Time to most recent common ancestor (tMRCA) of *Mycobacterium tuberculosis*– complex lineages in Botswana.

| **Lineage** | **tMRCA**<br>**(Posterior mean)** | **tMRCA**<br>**(Posterior median)** | **95% HPD interval** |
| --- | --- | --- | --- |
| L1 | 1727 | 1736 | 1607 – 1828 |
| L2 | 1900 | 1903 | 1854 – 1938 |
| L4.1.1 | 1729 | 1739 | 1587 – 1842 |
| L4.1.2 | 1695 | 1705 | 1546 – 1817 |
| L4.3.2 | 1941 | 1943 | 1911 – 1966 |
| L4.3.4 | 1808 | 1814 | 1723 – 1881 |
| L4.4 | 1739 | 1748 | 1621 – 1847 |
| L4.8 | 1853 | 1858 | 1782 – 1912 |

Abbreviations: HPD: highest posterior density.

**Figure 2.**
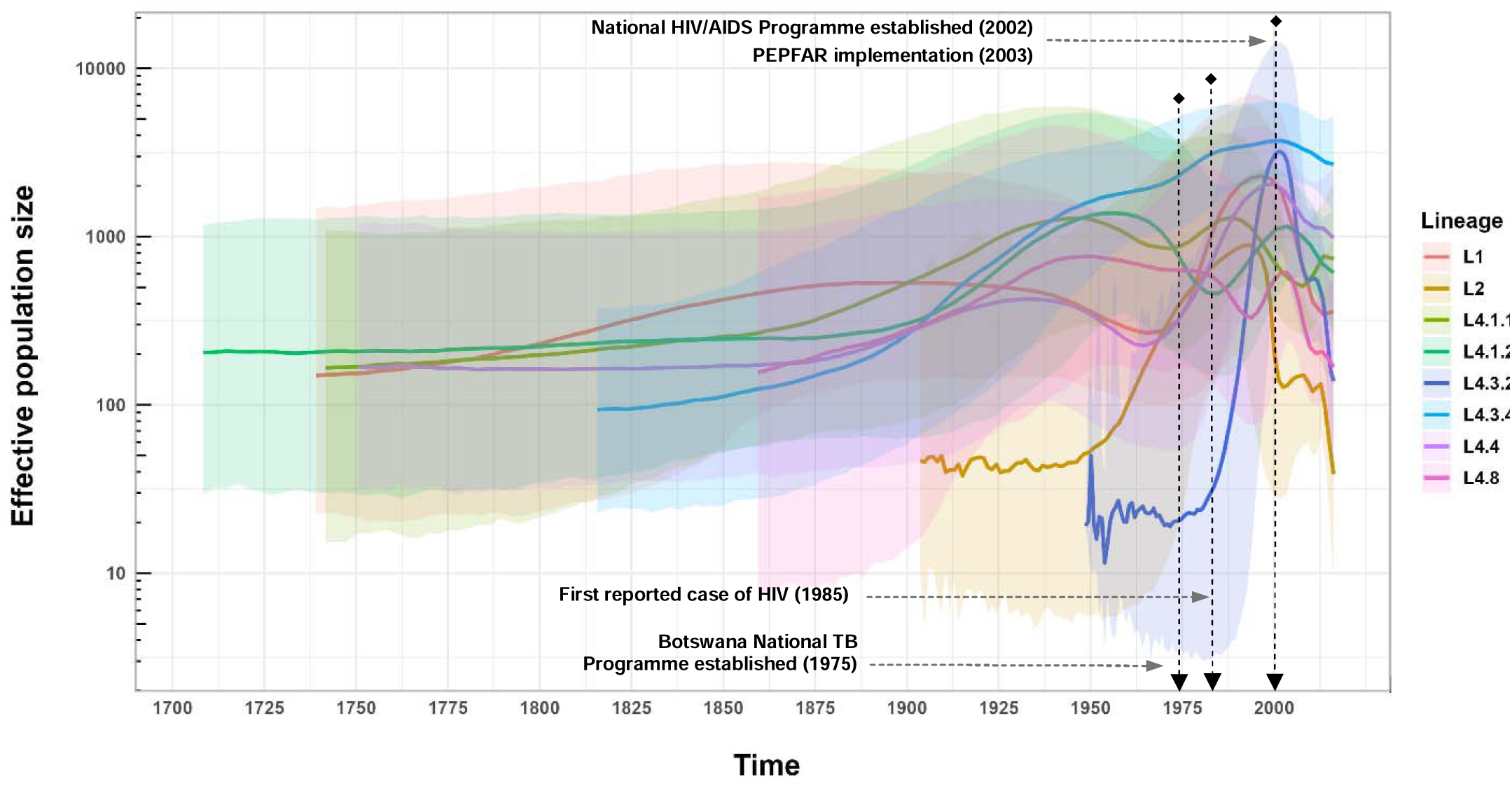
Inferred effective population size over time for *Mycobacterium tuberculosis–*complex lineages circulating in Botswana.

Using a 5-SNPs cutoff as an indicator of recent transmission, 48% of the isolates (652/1354) were clustered. The cluster sizes varied, ranging from 2 isolates per cluster to 23 isolates in the largest cluster. The median cluster size for each lineage was as follows: L1 and L2: 2 (range: 2–7), L4.1.1: 3 (range: 2–23), L4.1.2: 3 (range: 2–12), L4.3.2 and L4.4: 2 (range 2–8), L4.3.4: 2 (range: 2–19), and L4.8: 3.5 (range: 2–9). Compared to L1, we found that isolates belonging to any other lineages were more likely to be part of a cluster (Table 4). Isolates belonging to L2, L4.1.1, and L4.8 had the highest cluster proportion. Additionally, the cluster proportion for the predominant L4.3.4 was significantly higher in Ghanzi than Gaborone (57% vs. 40%; *p-*value <0.001; data not shown) These findings remained robust when we repeated the analysis using a 12-SNPs cutoff (Supplementary Table 2).

**Table 4.** Genomic cluster proportions (based on a 5-SNPs cutoff) of the *Mycobacterium tuberculosis*–complex lineages.

| Lineage | Cluster proportions | Crude OR (95% CI) | Adjusted<sup>1</sup> OR (95% CI) |
| --- | --- | --- | --- |
| L1 | 25/86 | 1 | 1 |
| L2 | 42/72 | 3.42 (1.78, 6.69) | 3.26 (1.68, 6.47) |
| L4.1.1 | 83/141 | 3.49 (1.99, 6.28) | 3.44 (1.94, 6.25) |
| L4.1.2 | 74/149 | 2.41 (1.38, 4.29) | 2.26 (1.28, 4.07) |
| L4.3.2 | 52/117 | 1.95 (1.09, 3.56) | 1.91 (1.05, 3.52) |
| L4.3.4 | 190/400 | 2.21 (1.35, 3.71) | 1.68 (1.00, 2.89) |
| L4.4 | 82/189 | 1.87 (1.09, 3.27) | 1.78 (1.03, 3.16) |
| L4.8 | 55/92 | 3.63 (1.96, 6.86) | 3.27 (1.74, 6.26) |

Abbreviations: SNP: single nucleotide polymorphism. OR: odds ratio. CI: confidence interval.
<sup>1</sup>Adjusted for age, gender, HIV status, and district.

## Discussion

Few studies to date have examined lineage-specific, long-term transmission dynamics of Mtbc strains in a high HIV/TB setting, largely due to the lack of population-level WGS datasets over extended sampling periods.^16, 21^ By reconstructing the evolutionary history and spread of Mtbc strains of different lineages using WGS collected over a four-year period, we observed varying transmission dynamics and demographic trajectories for extant lineages circulating in Botswana. The expansion of many lineages coincided with the onset of the HIV/AIDS epidemic, while their contraction aligned with the implementation of antituberculosis and antiretroviral treatment programs.

With the exception of strains of L4.1.1, L4.1.2 and L4.8, strains of most lineages experienced an expansion of various magnitude in the second half of the 20th century, correlating closely with the region’s high TB burden.^22^ Notably, this period coincides with the emergence and rapid escalation of the HIV epidemic. Botswana experienced one of the world’s most severe HIV epidemics, with initial cases emerging in the 1980s.^23^ A phylodynamic study of HIV dated the virus’s emergence in Botswana around 1960, followed by rapid growth until stabilization post-1990.^24^ HIV increases the susceptibility of TB disease and is the strongest risk factor for reactivating latent TB infections due to compromised immunity.^25^ Further, the HIV crisis was exacerbated by rapid urbanization driven by economic growth in the mining industry. A network analysis of micro-census data from Botswana between 1981 and 2011 suggests a highly mobile population with considerable movement between urban and rural areas.^26^ Mining towns, where HIV cases initially emerged, acted as major migration hubs for both inflow and outflow of populations. In Selebi Phikwe, a mining town for copper and nickel in Botswana, HIV prevalence had surged to 50% in 2000.^27^ These towns also became hubs of TB transmission due to the high-risk conditions associated with mining and the growing HIV prevalence.^28, 29^ As the HIV epidemic escalated, the number of individuals susceptible to Mtbc infection and reactivation increased, driving the expansion of Mtbc populations. Thus, the intersection of the HIV epidemic, mining-driven urbanization, and high population mobility likely played a critical role in the significant expansion of strains from most Mtbc lineages in the latter part of the 20th century. Nevertheless, this expansion was not observed across all lineages. It’s possible that Mtbc strains may exhibit varying capacities for TB transmission or reactivation influenced by their interactions with the host in the context of HIV-related immunosuppression.^15, 17, 30^ We observed a marked decline in Mtbc populations at the turn of the 21st century, which aligned with a consistent decrease in TB incidence from 2000 to 2016.^31^ Several nationwide programs likely contributed to the reduction. The most significant decrease in L1, L4.3.2, L4.3.4, L4.4, and L4.8 occurred beginning the early 2000s, coinciding with the launch of the Masa Antiretroviral Therapy (ART) Programme in 2002.^23^ This program, also known as the Masa Programme, initially aimed to provide free ART to individuals with advanced immune suppression but eligibility expanded over the years. “Masa” means “new dawn” in Setswana, symbolizing hope and a fresh start for those affected by the epidemic. Shortly after, the President’s Emergency Plan for AIDS Relief (PEPFAR) was launched, focusing on combating HIV/AIDS globally, with a significant emphasis on sub-Saharan Africa.^32^ Botswana, with its high HIV prevalence rates, was a key focus country for PEPFAR. Both initiatives aimed to support those affected by HIV/AIDS through comprehensive treatment, prevention, and care programs, thereby reducing HIV transmission.^23, 32, 33^ As a result, Botswana had significantly increased number of people receiving ART, reduced the rate of new HIV infections, and improved the overall healthcare system’s capacity to manage HIV/AIDS.^32, 33, 34^ While PEPFAR and Masa Programme primarily targeted the HIV/AIDS epidemic, their impact on reducing HIV transmission indirectly led to a significant reduction in the burden of TB in Botswana.

Our study highlighted significant heterogeneity in transmission dynamics among strains of different Mtbc lineages analysed. We found that L1 strains maintained a relatively stable effective population size until experiencing a significant expansion following the emergence of HIV. L1 is most prevalent in regions bordering the Indian Ocean, and several large-scale phylogeographic studies have shown that South Asia and eastern Africa played critical roles in dispersing this lineage via maritime trade.^35, 36^ It is possible that MRCA of L1 strains was introduced into Botswana through subsequent migrations between eastern and southern regions of the continent. Post establishment, evidence suggests that L1 likely expanded due to changing environmental conditions (*e.g.* increased population densities, malnutrition, immunosuppression), rather than from large migration or transmission events.^35^ Consistent with previous findings, our study found that L1 had the lowest proportion of cases that belonged to clusters.^14, 16, 37^ A phylodynamic study from Vietnam, an L1-endemic region, concluded that compared to L2 and L4, the disease burden due to L1 in the region was associated with reactivation of long-term remote infections, evidenced by larger SNP differences and limited inferred migration events among L1 isolates.^38^ Similarly, in Botswana, reactivation of latent TB infections likely contributes to new cases of L1 and may have driven this lineage’s expansion as population immunity waned due to HIV. L4.4 strains followed similar demographic trends but showed a slower decline in population size after nationwide introduction of ART interventions, which may be linked to its higher proportion of clustered cases due to recent transmission compared to L1 strains.

Notably, L2 strains underwent rapid expansion in effective population size shortly after their estimated emergence in the early 20th century. Phylogenetic studies suggest that L2.2, often associated with the Beijing genotype, likely originated in China.^21, 39^ The 19th and 20th centuries were characterized by intense interaction between Europe and China, driven by trade, imperial ambitions, and cultural exchanges. Following the Opium Wars, numerous Chinese cities became treaty ports where European powers had extraterritorial rights and conducted trade with relative freedom. Prominent treaty ports included Shanghai, Guangzhou, and Tianjin, which became hubs of international trade and cultural exchange. Additionally, during the late 19th and early 20th centuries, Chinese laborers were often recruited for mining and railway construction projects worldwide, including in Africa.^40^ South Africa, which borders Botswana, had a significant number of Chinese laborers working in its mines.^51^ This movement could have facilitated the introduction and spread of L2.2 (with putative origins in Beijing, China) to southern Africa.^50^ L2 has been associated with hypervirulence (ability to cause disease shortly after infection) and high transmissibility, supported by both clinical evidence and animal experiments.^41, 42^ In our study, we also observed that L2 had one of the highest proportions of genomic clustering due to recent transmission. However, L2 strains experienced a decline in the early 1990s before the implementation of country-wide ART programs. This decline might reflect a combination of factors, including implementation of facility-based directly observed therapy, local pathogen-host dynamics and its challenges in competing with other well-adapted indigenous lineages.^15^ The discrepancy between high genotypic clustering proportion and population dynamic of L2 strains in Botswana highlights that genotypic clustering analysis alone does not necessarily capture the dynamics of an epidemic, nor does it always correlate with the population size of a specific Mtbc lineage. Our finding also contrasts sharply with the phylodynamic study in Vietnam, where L2 showed clear patterns of overtaking endemic L1 strains over time.^38^ Overall, these findings imply that the competitive dynamics between strains of different lineages can vary significantly depending on the region and the specific lineages involved, and that studying lineage features without accounting for geographic or host genetic background can obscure important host-pathogen interactions.^15, 43^

Our analysis indicates that most TB cases in Botswana were caused by L4 strains, specifically L4.3 strains. This finding aligns with previous reports of L4-belonging Latin American and Mediterranean sublineage’s dominant presence in southern Africa region.^44, 45, 46, 47^ Based on the sampled genetic diversity, we estimated that most L4 sublineages trace back to MRCAs that emerged between the late 17th and 19th centuries. Recent genomic studies of Mtbc suggests that the introduction and spread of L4 coincide with historical events involving European colonization and subsequent socioeconomic changes.^44, 48^ European colonization of South Africa, Botswana’s neighboring country, began in earnest with the establishment of a Dutch settlement at the Cape of Good Hope in 1652.^49^ Later, British colonization intensified in early 19th century, and Botswana (then Bechuanaland) became a British protectorate in 1885. The establishment of colonial administration and subsequent movements of European settlers between Europe, South Africa and Botswana during this period may have facilitated the establishment and spread of L4 strains still observed today.

The most common Mtbc lineage found in Botswana was L4.3.4, and it was overwhelmingly more prevalent in Ghanzi than Gaborone (67% vs. 21%). Another study using a different genotyping technique noted this, but was unable to differentiate whether it was due to recent transmission or reactivation of latent infections.^50^ The effective population size of L4.3.4 strains grew steadily since its introduction around 1880 and only began to decline slightly after country-wide ART programs began. Unlike other lineages, L4.3.4’s expansion during the HIV epidemic was modest. This contrast may be explained by the higher prevalence of this lineage in Ghanzi, which has lower HIV prevalence compared to Gaborone. Prior to the study, approximately 36% of TB patients in Ghanzi were coinfected with HIV, compared to 70% in Gaborone,^51^ suggesting a less pronounced impact of the HIV epidemic on TB transmission dynamics in Ghanzi. In our analysis, L4.3.4 displayed moderate clustering proportions due to recent transmission, falling between the lowest (L1) and the highest groups (e.g., L2, L4.1.1, L4.8). However, it exhibited significantly higher cluster proportions in Ghanzi compared to Gaborone. Taken collectively, we propose that the success of L4.3.4 is driven primarily by sustained ongoing transmission, rather than reactivation of latent infections.^50^ This highlights the need for public health efforts to prioritize activities that interrupt transmission, such as enhancing contact tracing,^52^ improving infection control measures,^53^ and ensuring timely diagnosis and treatment to reduce disease burden, especially in a rural setting like Ghanzi.^54^

L4.3.4 displayed clear competitive advantage as evidenced by its dominant presence in both districts despite not being the earliest lineage to emerge in Botswana. It’s also interesting to observe that L4.3.2, a strain closely related to L4.3.4, emerged around the time of estimated HIV introduction in Botswana.^24^ However, unlike L4.3.4’s steady growth, L4.3.2 experienced a rapid expansion followed by a sharp decline, mirroring remarkably well with the timing of the HIV epidemic. The striking difference in population dynamics between two closely related strains warrant further investigation to determine whether strain-specific mutations contribute to distinct phenotypes, and/or if potential host-pathogen interactions influence their respective trajectories and the success of L4.3.4.

L4.1.1, L4.1.2, and L4.8 strains share similar demographic trends with large overlapping CIs. Notably, L4.1.1 and L4.8 exhibited the highest cluster proportions due to recent transmission, almost doubling the estimate compared to L4.3.4. This variation in genotypic clustering within L4 sublineages was also observed in San Francisco, USA,^55^ underscoring sublineage-level heterogeneity and the necessity of investigating them individually. Furthermore, L4.1.1 was the only lineage that expanded after the implementation of country-wide ART programs. In addition to possible sublineage-specific characteristics (*e.g.* virulence), other socioecological drivers, for example, overcrowding or malnutrition, may be responsible for the expansion and contraction of this sublineage.^56^ However, the relatively wide CIs associated with the effective population size of these sublineages limit our ability to analyze local features of their curves.^57^

Our study had limitations. First, the available genetic data may not fully represent the diversity of Mtbc strains circulating in Botswana. However, the greater Gaborone and Ghanzi district encompass a considerable segment (20%–25%) of the total population, and sampling in Gaborone likely captured a substantial portion of Mtbc strain diversity due to its role as a central urban hub for inflow and outflow of population. Second, Mtbc isolates were limited to sputa. It is possible that some lineages may be under-represented because of the predication for pulmonary clinical presentation.^58^ Third, coalescent models (*e.g.* Skyride model applied in our analysis) use pathogen genomic data to estimate the timescale of population dynamics.^59^ The relatively low mutation rate of Mtbc can result in broad uncertainty intervals for estimated effective population sizes and divergence times, making it challenging to pinpoint precise timings for events such as introductions or expansions of lineages. Inclusion of Mtbc sequences as they become available in the future may improve the reconstruction of Mtbc population dynamics and reduce uncertainty associated with population parameters. Lastly, we acknowledge the limitation of using a fixed-SNP threshold to determine transmission clusters.^60^ The chosen threshold is often arbitrary and might not accurately reflect recent transmission. However, a SNP cutoff provides a standardized approach, making it easier to compare results across different studies and useful for public health investigations. The chosen 5-SNP cutoff for determining TB transmission clusters is common in molecular epidemiology and has been shown to provide an informative overview of local transmission patterns.^61, 62^

In summary, our study provides insight into the recent evolutionary origins and population dynamics of strains of different Mtbc lineages in Botswana. We highlight the heterogeneous transmission dynamics of key lineages circulating in Botswana and emphasize that increasing awareness of these heterogeneities may be essential to further reducing TB burden. Similar to viral pathogens, we recommend routine molecular surveillance and phylodynamic analysis to monitor local TB transmission. This could enable public health programs to effectively track the spread of different Mtbc strains, identify those that are expanding over time, and tailor control strategies based on the epidemiological trends of circulating Mtbc strains within a setting.

## Methods

### Data source and study setting

Data were obtained from a population-based TB study conducted between 2012 and 2016. Study procedures have been described in detail elsewhere.^20^ Briefly, the study enrolled individuals of all ages with presumed TB in the greater Gaborone and Ghanzi districts of Botswana. Gaborone, located in the southeast and bordering South Africa, is the capital city with a high population density of 1,370 persons/km^2^ in 2011. In contrast, Ghanzi is a rural district located in western Botswana with the lowest population density in the country according to the 2011 census, at 0.37 persons/km^2^.^63^ Despite Ghanzi’s low population density, it consistently has the highest TB incidence in the country. Prior to the study, the TB incident rate per 100,000 people in Botswana, and Gaborone and Ghanzi districts were approximately 445, 440, and 722 cases, respectively.^64^

During the enrollment process, participants without a documented HIV status or with negative test results over 12 months old were offered rapid HIV testing. Each participant was required to provide at least one expectorated sputum sample. Sputum induction with nebulized hypertonic saline solution was performed for those who had difficulty producing sputum spontaneously. The Kopanyo Study was approved by the U.S. Centers for Disease Control and Prevention, the Botswana Ministry of Health and Wellness, and the University of Pennsylvania Institutional Review Boards. Additionally, approval of this secondary data analysis was obtained from University of California at Irvine Institutional Review Board. Written informed consent was obtained from all participants prior to enrollment.

### Culture and DNA extraction

The sputum specimens underwent decontamination using the N-acetyl-L-cysteine and sodium hydroxide method. After processing, the specimens were inoculated into Mycobacteria Grow Indicator Tubes (MGIT) 960 system (Becton Dickinson Microbiology Systems, Sparks, MD, USA) at the Botswana national reference laboratory in Gaborone. Weekly monitoring of mycobacteria growth occurred, and if no growth was observed within 8 weeks, it was classified as culture negative. For culture positive colonies, Mtbc DNA was extracted with GenoLyse kit (Hain Lifescience, Germany) following the manufacturer’s protocol. Genomic extracts were stored at −80°C, and those containing a sufficient amount of DNA (at least 0.05ng/uL) were sent to the Research Center Borstel for WGS.

### Whole genome sequencing and bioinformatics

Genomic libraries were prepared with Illumina Nextera XT kit to generate 2×150bp paired-end reads for sequencing using the Illumina NextSeq 500 platform. An automated bioinformatics pipeline, MTBseq, was used for WGS data analysis.^65^ MTBseq combined all necessary steps needed for analysis of WGS data. Briefly, raw sequences were aligned to the pan-susceptible, H37Rv reference genome (GenBank NC000962.2) with BWA-mem software and Samtools. Next, base call recalibration and realignment of reads around insertions and deletions was performed with GATKv3. Variant calling was performed using Samtools mpileup and in-house scripts, and positions were considered reliable with 75% allele frequency, at least 4 calls with a phred score of 20 or more, and a minimum of 4 reads mapped in each direction (forward and reverse orientation). Libraries were considered as high quality and further analyzed when at least 50x coverage was achieved and 95% of the reference genome covered. Next, genomes were annotated with known associations to antibiotic resistance. Finally, phylogenetically informative SNPs based on existing literature were used for lineage classification of the input samples,^13^ excluding genes associated with antibiotic resistance, insertions and deletions, repetitive regions (PPE and PE-PGRS gene families), and consecutive variants in a 12bp window.

### Phylogenetic reconstruction

We extracted variable sites from whole genome alignments of Mtbc isolates to create the fasta files used for phylogenetic analysis. To illustrate the overall population structure of Mtbc lineages in Botswana, we used IQ-TREE v1.6.12 to reconstruct maximum likelihood phylogenies for each major Mtbc lineage, with isolates possibly containing mixed Mtbc strains excluded from the dataset.^66^ The Hasegawa, Kishino, and Yano (HKY) model of nucleotide evolution was assumed,^67^ and SNP ascertainment bias correction was specified. Additionally, time-measured phylogenetic trees for specific lineages included in the coalescent-based analysis were estimated using Bayesian method with details described below.

### Coalescent-based demographic reconstruction

We reconstructed Mtbc population dynamics for L1, L2, and L4 sublineages that had at least 70 samples using BEAST v1.10.4.^68^ The HKY nucleotide substitution model with gamma-distributed rate heterogeneity (discretized to 4 categories) was used.^67^ We specified a strict molecular clock model, with the substitution rate assumed to be distributed lognormal with a mean of 1 and a sigma of 1.25. This prior translates to 95% of the probability mass between 0.09 and 4.2 SNPs per site per year (s/s/y). We also explored alternative priors more aligned with the published substitution rate of Mtbc,^69^ such as a lognormal distribution with a mean of 0.001 and a sigma of 2 (i.e. 95% of probability mass between 2.4E-10 and 0.001 s/s/y), as well as a uniform distribution with upper and lower bounds set to 5E-7 and 1E-8 s/s/y, respectively. However, the posterior clock rate remained robust to these different prior specifications. To estimate the mycobacterial population size change through time, we adopted the coalescent-based Gaussian Markov random fields Bayesian Skyride tree prior.^59^ This approach offers greater flexibility compared to the Bayesian Skyline prior and does not require strong prior assumptions on the number of population size change points. Furthermore, to account for ascertainment bias, we made manual adjustments in the xml file to incorporate the number of invariant sites. This correction was applied individually for each xml file corresponding to a specific lineage. The Markov chain Monte Carlo (MCMC) chain for each lineage ran for 500 million iterations, with parameters and trees sampled every 50,000th iteration. We verified adequate mixing and posterior convergence using Tracer v1.7.2 after discarding the first 10% as burn-in. For a parameter to be considered sufficiently sampled, we aimed for an effective sample size of at least 200. If any parameter fell short of this threshold, we ran a second chain of 500 million iterations and combined the chains using LogCombiner v1.10.4. We note that L4.3.4 – the largest sublineage in our dataset – was the only one that required a second MCMC chain of 500 million iterations to achieve sufficient mixing and sampling. Finally, a consensus tree summarizing the sampled posterior trees for each lineage was constructed using the maximum clade credibility method.

### Genomic clustering analysis

We explored differences in patterns of recent transmission between Mtbc lineages that were included in the coalescent analysis. Genetically similar Mtbc strains under a pre-specified SNP threshold (usually between 5 and 12-SNPs) are interpreted as a consequence of recent transmission events^61^. We assigned cases into clusters, which was defined as two or more individuals with Mtbc strains sharing ≤5-SNPs differences, as determined by pairwise SNP comparison.^61, 62^ Next, we regressed clustered or non-clustered case on Mtbc lineage in a logistic model, adjusting for potential confounding by age, gender, enrollment district, and HIV status. To evaluate the robustness of our findings, we also conducted logistic regression analysis using a 12-SNPs cutoff for identifying clustered cases. All statistical analysis and phylogenetic tree visualizations were conducted in R version 4.2.0.^70^ WGS data from this study has been submitted to the European Nucleotide Archive (ENA) under study accession PRJEB:62480.

## Data Availability

All data produced in the present study are available upon reasonable request to the authors

## Acknowledgements

We would like to thank the participants who made this research possible.

## Author contributions

These authors contributed equally: Sanghyuk S. Shin and Stefan Neimann.

Q.W. contributed to project conception, performed the data analysis, and drafted the manuscript.

I.B. and S.N. processed the whole genome sequencing data and performed the bioinformatic analysis. C.M., N.Z., P.K.M., A.F., R.B., J.E.O., and T.L.M. contributed to the Kopanyo Study design, participant recruitment, data collection, and quality control. V.M.M., S.S.S., and T.F.B. contributed to project conception, supervised data analysis and interpretation of results. All authors were involved in revising the manuscript and approved the final version.

## References

1. World Health Organization. Global Tuberculosis Report 2023. World Health Organization (2023).

2. Walensky, R. P., Walke H. T., Fauci A. S. SARS-CoV-2 variants of concern in the United States-challenges and opportunities. JAMA 325, 1037–1038 (2021).

3. Kinganda-Lusamaki, E., et al. Integration of genomic sequencing into the response to the Ebola virus outbreak in Nord Kivu, Democratic Republic of the Congo. Nature Medicine 27, 710–716 (2021).

4. Grubaugh, N. D., et al. Genomic epidemiology reveals multiple introductions of Zika virus into the United States. Nature 546, 401–405 (2017).

5. Yuan, D., et al. HIV-1 genetic transmission networks among people living with HIV/AIDS in Sichuan, China: a genomic and spatial epidemiological analysis. Lancet Reg Health West Pac 18, 100318 (2022).

6. Skaathun, B., et al. HIV-1 transmission dynamics among people who inject drugs on the US/Mexico border during the COVID-19 pandemic: a prosepective cohort study. Lancet Reg Health Am 33, 100751 (2024).

7. Attwood, S. W., Hill S. C., Aanensen D. M., Connor T. R., Pybus O. G. Phylogenetic and phylodynamic approaches to understanding and combating the early SARS-CoV-2 pandemic. Nat Rev Genet 23, 547–562 (2022).

8. Rife, B. D., et al. Phylodynamic applications in 21(st) century global infectious disease research. Glob Health Res Policy 2, 13 (2017).

9. Bos, K. I., et al. Pre-Columbian mycobacterial genomes reveal seals as a source of New World human tuberculosis. Nature 514, 494–497 (2014).

10. Comas, I., et al. Out-of-Africa migration and Neolithic coexpansion of Mycobacterium tuberculosis with modern humans. Nat Genet 45, 1176–1182 (2013).

11. Coscolla, M., et al. Phylogenomics of Mycobacterium africanum reveals a new lineage and a complex evolutionary history. Microb Genom 7, (2021).

12. Ngabonziza, J. C. S., et al. A sister lineage of the Mycobacterium tuberculosis complex discovered in the African Great Lakes region. Nat Commun 11, 2917 (2020).

13. Coll, F., et al. A robust SNP barcode for typing Mycobacterium tuberculosis complex strains. Nat Commun 5, 4812 (2014).

14. Freschi, L., et al. Population structure, biogeography and transmissibility of Mycobacterium tuberculosis. Nat Commun 12, 6099 (2021).

15. Gagneux, S. Host-pathogen coevolution in human tuberculosis. Philosophical transactions of the Royal Society of London Series B, Biological sciences 367, 850–859 (2012).

16. Guerra-Assuncao, J. A., et al. Large-scale whole genome sequencing of M. tuberculosis provides insights into transmission in a high prevalence area. Elife 4, (2015).

17. Coscolla, M., Gagneux S. Consequences of genomic diversity in Mycobacterium tuberculosis. Semin Immunol 26, 431–444 (2014).

18. Mathema, B., et al. Epidemiologic consequences of microvariation in Mycobacterium tuberculosis. J Infect Dis 205, 964–974 (2012).

19. Gagneux, S., et al. Variable host-pathogen compatibility in Mycobacterium tuberculosis. Proc Natl Acad Sci U S A 103, 2869–2873 (2006).

20. Zetola, N. M., et al. Protocol for a population-based molecular epidemiology study of tuberculosis transmission in a high HIV-burden setting: the Botswana Kopanyo study. BMJ open 6, e010046 (2016).

21. Liu, Q., et al. China’s tuberculosis epidemic stems from historical expansion of four strains of Mycobacterium tuberculosis. Nat Ecol Evol 2, 1982–1992 (2018).

22. Dodd, P. J., et al. Transmission modeling to infer tuberculosis incidence prevalence and mortality in settings with generalized HIV epidemics. Nat Commun 14, 1639 (2023).

23. Ramogola-Masire, D., et al. Botswana’s HIV response: Policies, context, and future directions. J Community Psychol 48, 1066–1070 (2020).

24. Wilkinson, E., Engelbrecht S., de Oliveira T. History and origin of the HIV-1 subtype C epidemic in South Africa and the greater southern African region. Sci Rep 5, 16897 (2015).

25. Corbett, E. L., et al. The growing burden of tuberculosis: global trends and interactions with the HIV epidemic. Archives of internal medicine 163, 1009–1021 (2003).

26. Song, J., et al. Population mobility and the development of Botswana’s generalized HIV epidemic: a network analysis. medRxiv, (2023).

27. UNAIDS. Botswana: Country Factsheets. UNAIDS (2004).

28. Stuckler, D., Basu S., McKee M., Lurie M. Mining and risk of tuberculosis in sub-Saharan Africa. Am J Public Health 101, 524–530 (2011).

29. Basu, S., Stuckler D., Gonsalves G., Lurie M. The production of consumption: addressing the impact of mineral mining on tuberculosis in southern Africa. Global Health 5, 11 (2009).

30. Fenner, L., et al. HIV infection disrupts the sympatric host-pathogen relationship in human tuberculosis. PLoS Genet 9, e1003318 (2013).

31. The World Bank. Incidence of tuberculosis in Botswana. The World Bank Group (2022).

32. Chin, R. J., Sangmanee D., Piergallini L. PEPFAR funding and reduction in HIV infection rates in 12 focus sub-Saharan African countries: a quantitative analysis. Int J MCH AIDS 3, 150–158 (2015).

33. Farahani, M., et al. Outcomes of the Botswana national HIV/AIDS treatment programme from 2002 to 2010: a longitudinal analysis. Lancet Glob Health 2, e44–50 (2014).

34. Mine, M., et al. Progress towards the UNAIDS 95-95-95 targets in the Fifth Botswana AIDS Impact Survey (BAIS V 2021): a nationally representative survey. Lancet HIV 11, e245–e254 (2024).

35. O’Neill, M. B., et al. Lineage specific histories of Mycobacterium tuberculosis dispersal in Africa and Eurasia. Mol Ecol 28, 3241–3256 (2019).

36. Menardo, F., et al. Local adaptation in populations of Mycobacterium tuberculosis endemic to the Indian Ocean Rim. F1000Res 10, 60 (2021).

37. Couvin, D., Reynaud Y., Rastogi N. Two tales: Worldwide distribution of Central Asian (CAS) versus ancestral East-African Indian (EAI) lineages of Mycobacterium tuberculosis underlines a remarkable cleavage for phylogeographical, epidemiological and demographical characteristics. PLoS One 14, e0219706 (2019).

38. Holt, K. E., et al. Frequent transmission of the Mycobacterium tuberculosis Beijing lineage and positive selection for the EsxW Beijing variant in Vietnam. Nat Genet 50, 849–856 (2018).

39. Merker, M., et al. Evolutionary history and global spread of the Mycobacterium tuberculosis Beijing lineage. Nat Genet 47, 242–249 (2015).

40. Altan, S. Chinese Workers of the World: Colonialism, Chinese Labor, and the Yunnan– Indochina Railway. Stanford University Press (2024).

41. Hanekom, M., et al. Mycobacterium tuberculosis Beijing genotype: a template for success. Tuberculosis (Edinb) 91, 510–523 (2011).

42. Parwati, I., van Crevel R., van Soolingen D. Possible underlying mechanisms for successful emergence of the Mycobacterium tuberculosis Beijing genotype strains. The Lancet Infectious diseases 10, 103–111 (2010).

43. Gagneux, S. Ecology and evolution of Mycobacterium tuberculosis. Nat Rev Microbiol 16, 202–213 (2018).

44. Stucki, D., et al. Mycobacterium tuberculosis lineage 4 comprises globally distributed and geographically restricted sublineages. Nat Genet 48, 1535–1543 (2016).

45. Mogashoa, T., et al. Genetic diversity of Mycobacterium tuberculosis strains circulating in Botswana. PLoS One 14, e0216306 (2019).

46. Chihota, V. N., et al. Geospatial distribution of Mycobacterium tuberculosis genotypes in Africa. PLoS One 13, e0200632 (2018).

47. Viegas, S. O., et al. Molecular diversity of Mycobacterium tuberculosis isolates from patients with pulmonary tuberculosis in Mozambique. BMC Microbiol 10, 195 (2010).

48. Brynildsrud, O. B., et al. Global expansion of Mycobacterium tuberculosis lineage 4 shaped by colonial migration and local adaptation. Sci Adv 4, eaat5869 (2018).

49. Oliver, E., Oliver W. H. The colonisation of South Africa: a unique case. HTS Theological Studies 73, 1–8 (2017).

50. Click, E. S., et al. Phylogenetic diversity of Mycobacterium tuberculosis in two geographically distinct locations in Botswana - The Kopanyo Study. Infect Genet Evol 81, 104232 (2020).

51. Statistics Botswana. Botswana AIDS Impact Survey IV 2013 (BAIS IV): Report. National AIDS & Health Promotion Agency (2013).

52. Moonan, P. K., et al. A neighbor-based approach to identify tuberculosis exposure, the Kopanyo study. Emerging infectious diseases 26, 1010–1013 (2020).

53. Smith, J. P., et al. High-resolution characterization of nosocomial Mycobacterium tuberculosis transmission events in Botswana. Am J Epidemiol 192, 503–506 (2023).

54. Smith, J. P., et al. Characterizing tuberculosis transmission dynamics in high-burden urban and rural settings. Sci Rep 12, 6780 (2022).

55. Anderson, J., et al. Sublineages of lineage 4 (Euro-American) Mycobacterium tuberculosis differ in genotypic clustering. Int J Tuberc Lung Dis 17, 885–891 (2013).

56. Lopez, M. G., et al. Deciphering the tangible spatio-temporal spread of a 25-year tuberculosis outbreak boosted by social determinants. Microbiol Spectr 11, e0282622 (2023).

57. Du, D. H., et al. The effect of M. tuberculosis lineage on clinical phenotype. PLOS Glob Public Health 3, e0001788 (2023).

58. Click, E. S., Moonan P. K., Winston C. A., Cowan L. S., Oeltmann J. E. Relationship between Mycobacterium tuberculosis phylogenetic lineage and clinical site of tuberculosis. Clin Infect Dis 54, 211–219 (2012).

59. Minin, V. N., Bloomquist E. W., Suchard M. A. Smooth skyride through a rough skyline: Bayesian coalescent-based inference of population dynamics. Mol Biol Evol 25, 1459–1471 (2008).

60. Hatherell, H. A., et al. Interpreting whole genome sequencing for investigating tuberculosis transmission: a systematic review. BMC medicine 14, 21 (2016).

61. Walker, T. M., et al. Whole-genome sequencing to delineate Mycobacterium tuberculosis outbreaks: a retrospective observational study. The Lancet Infectious diseases 13, 137–146 (2013).

62. Zhang, X., et al. Exploring programmatic indicators of tuberculosis control that incorporate routine Mycobacterium tuberculosis sequencing in low incidence settings: a comprehensive (2017-2021) patient cohort analysis. Lancet Reg Health West Pac 41, 100910 (2023).

63. Statistics Botswana. Botswana Population and Housing Census. Statistics Botswana (2011).

64. Zetola, N. M., et al. Population-based geospatial and molecular epidemiologic study of tuberculosis transmission dynamics, Botswana, 2012–2016. Emerging infectious diseases 27, 835–844 (2021).

65. Kohl, T. A., et al. MTBseq: a comprehensive pipeline for whole genome sequence analysis of Mycobacterium tuberculosis complex isolates. PeerJ 6, e5895 (2018).

66. Nguyen, L. T., Schmidt H. A., von Haeseler A., Minh B. Q. IQ-TREE: a fast and effective stochastic algorithm for estimating maximum-likelihood phylogenies. Mol Biol Evol 32, 268–274 (2015).

67. Hasegawa, M., Kishino H., Yano T. Dating of the human-ape splitting by a molecular clock of mitochondrial DNA. J Mol Evol 22, 160–174 (1985).

68. Suchard, M. A., et al. Bayesian phylogenetic and phylodynamic data integration using BEAST 1.10. Virus Evol 4, vey016 (2018).

69. Menardo, F., Duchene S., Brites D., Gagneux S. The molecular clock of Mycobacterium tuberculosis. PLoS Pathog 15, e1008067 (2019).

70. R Core Team. R: a language and environment for statistical computing. R Foundation for Statistical Computing.

