## Supplementary information for "Phylodynamic analysis reveals disparate transmission dynamics of *Mycobacterium tuberculosis*-complex lineages in Botswana"

**Supplementary Appendix**

**Supplementary Figure 1.** Individual *Mycobacterium tuberculosis* complex lineage demographic trajectories

**Supplementary Figures 2a-h.** Time-measured, maximum clade credibility trees for individual *Mycobacterium tuberculosis* complex lineage

**Supplementary Table 1.** Posterior clock rate of the *Mycobacterium tuberculosis* complex lineages

**Supplementary Table 2.** Genomic cluster proportions (based on a 12-SNPs cutoff) of the *Mycobacterium tuberculosis* complex lineages

Supplementary Figure 1: Inferred effective population size over time for individual *Mycobacterium tuberculosis* complex lineage in Botswana

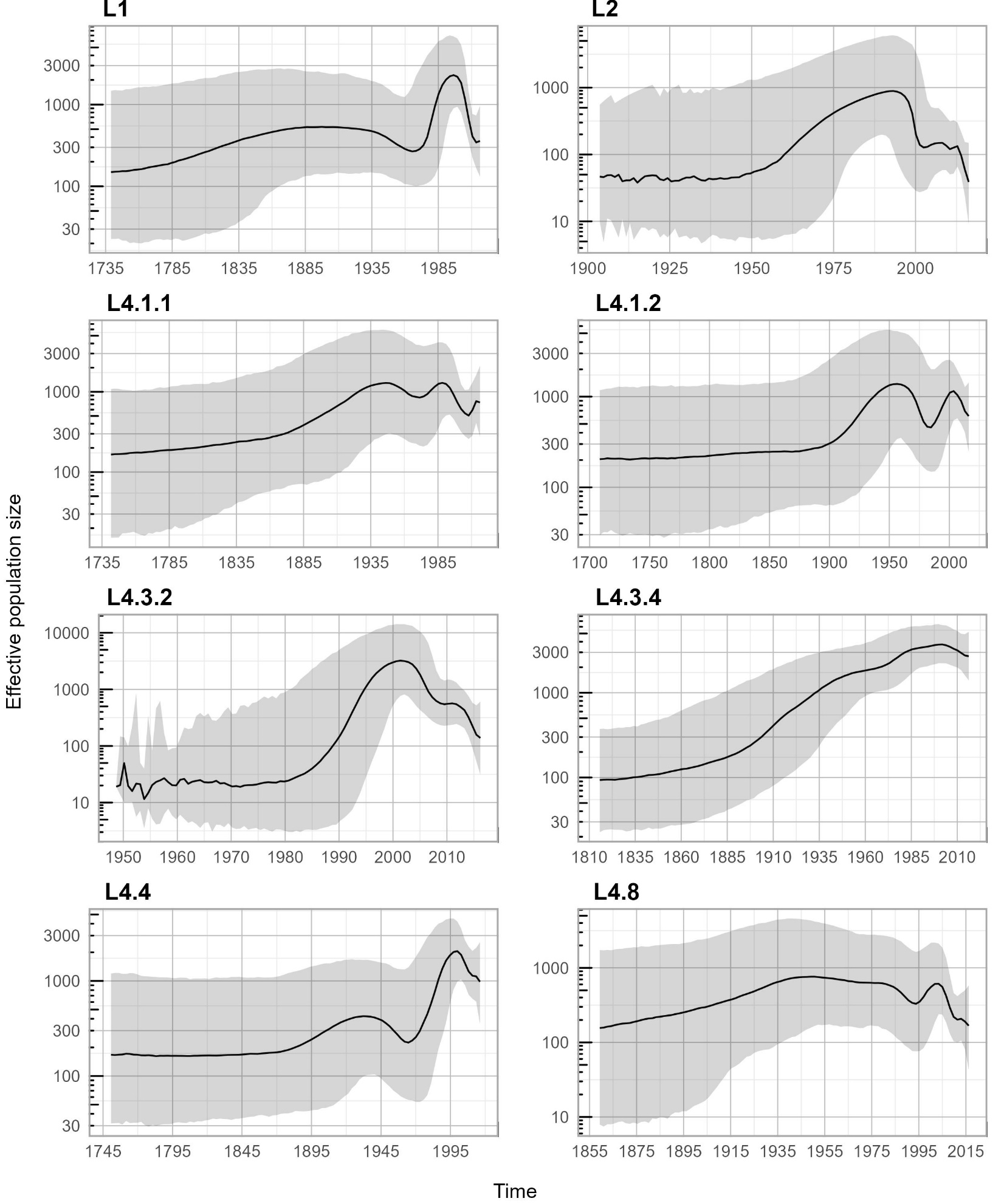

Supplementary Figure 2a: Maximum clade credibility tree of *Mycobacterium tuberculosis* complex lineage 1 with 95% highest posterior density intervals of node heights

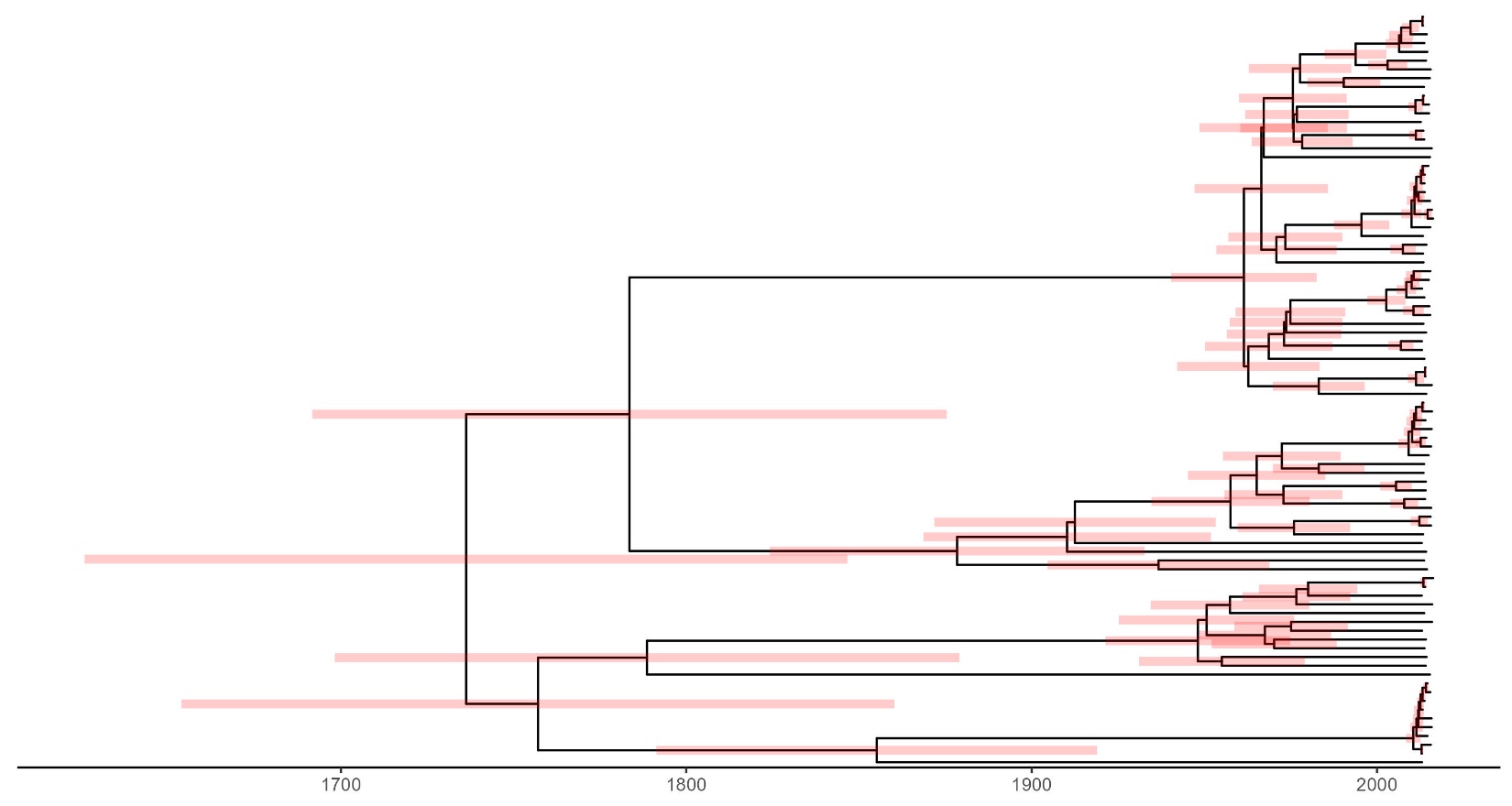

Supplementary Figure 2b: Maximum clade credibility tree of *Mycobacterium tuberculosis* complex lineage 2 with 95% highest posterior density intervals of node heights
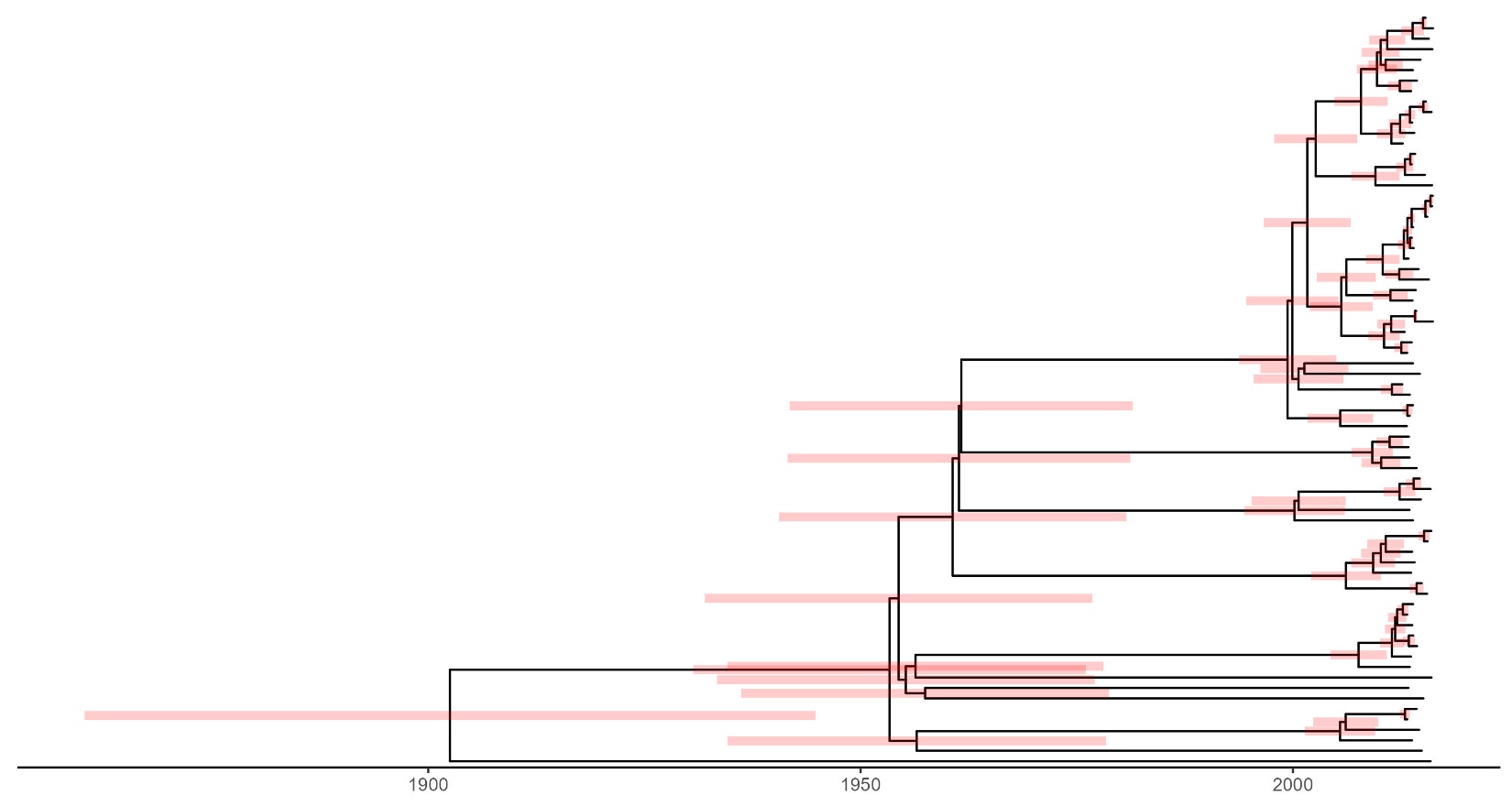

Supplementary Figure 2c: Maximum clade credibility tree of *Mycobacterium tuberculosis* complex lineage 4.1.1 with 95% highest posterior density intervals of node heights
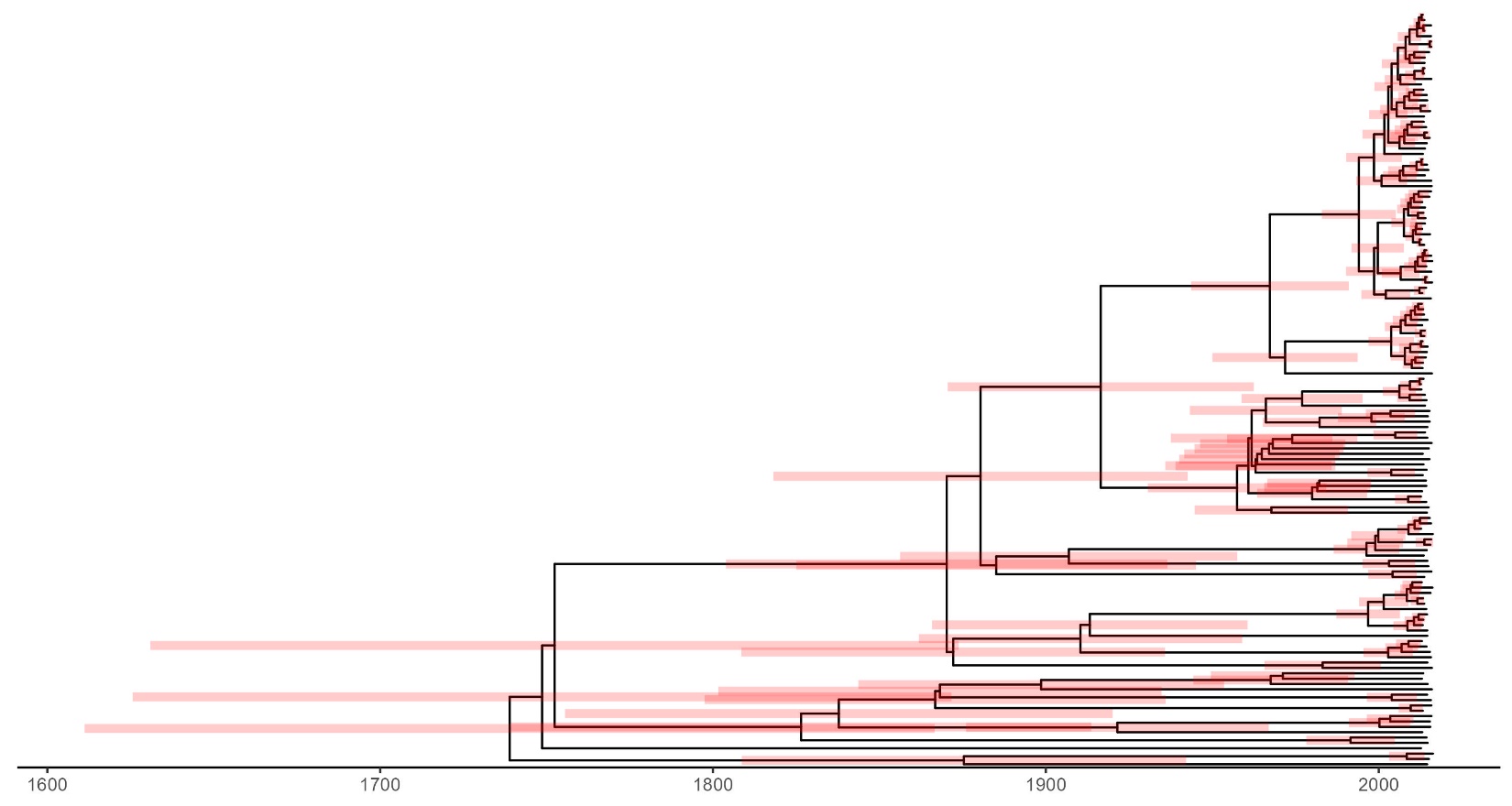

Supplementary Figure 2d: Maximum clade credibility tree of *Mycobacterium tuberculosis* complex lineage 4.1.2 with 95% highest posterior density intervals of node heights
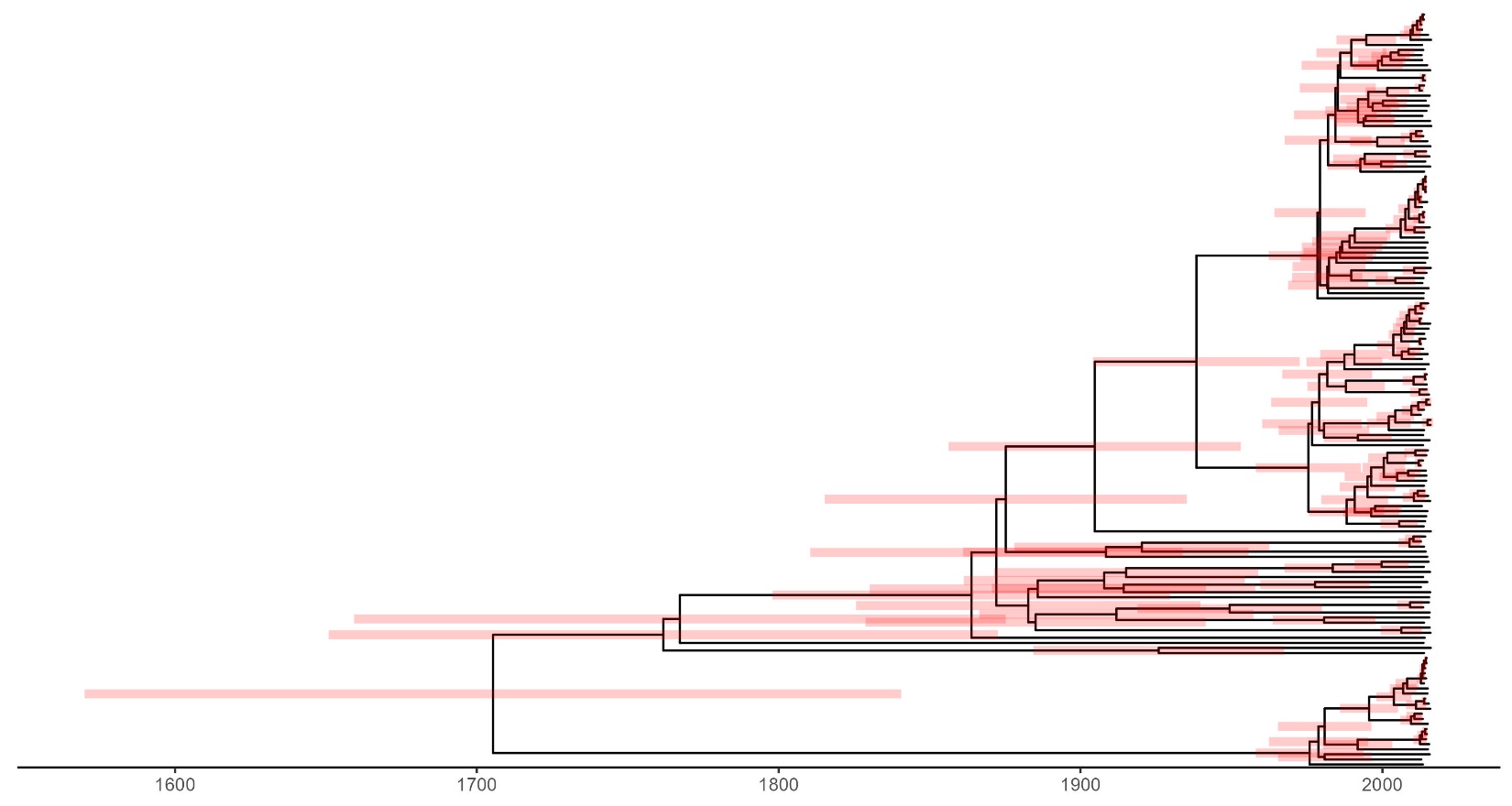

Supplementary Figure 2e: Maximum clade credibility tree of *Mycobacterium tuberculosis* complex lineage 4.3.2 with 95% highest posterior density intervals of node heights
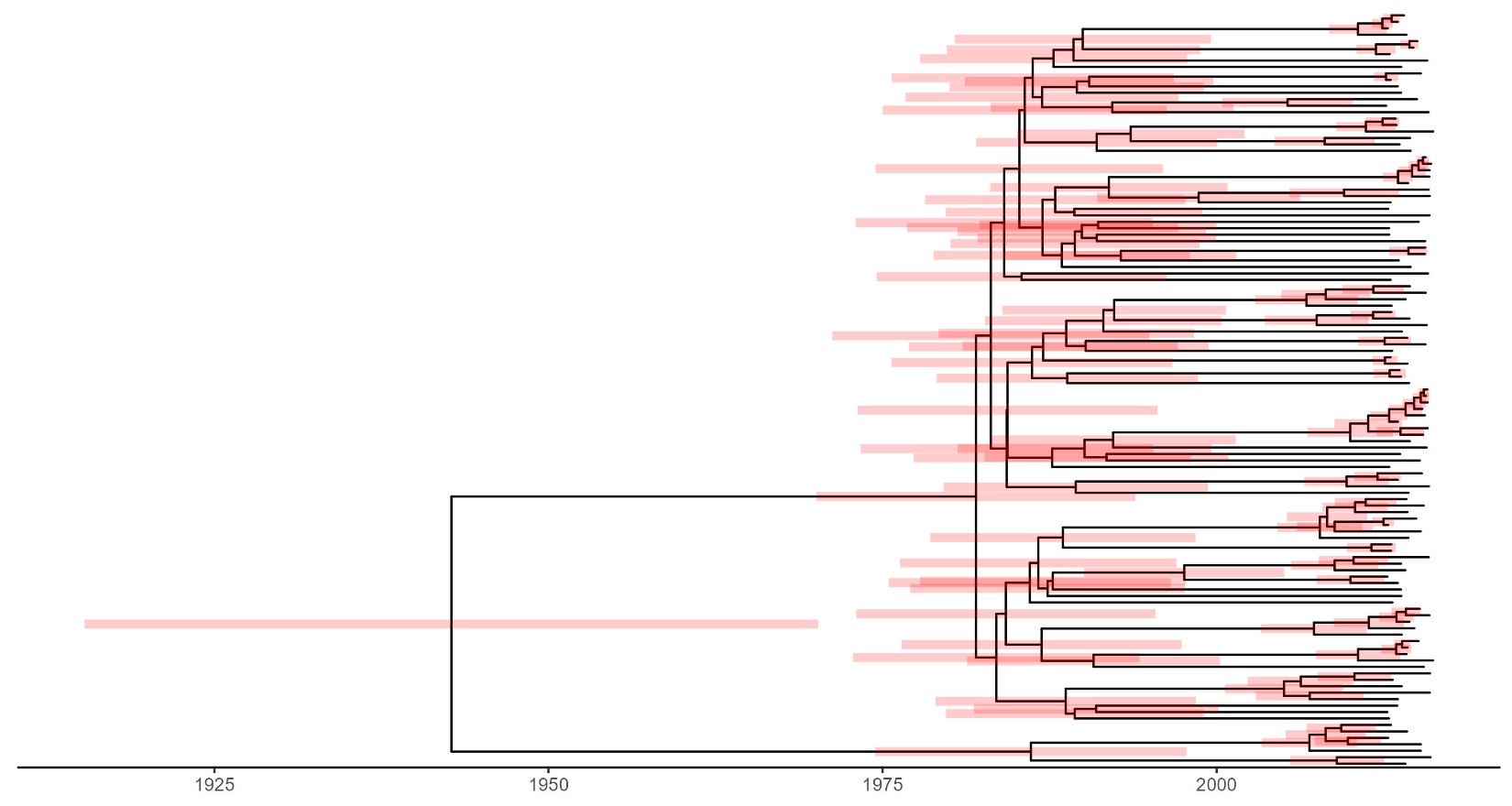

Supplementary Figure 2f: Maximum clade credibility tree of *Mycobacterium tuberculosis* complex lineage 4.3.4 with 95% highest posterior density intervals of node heights

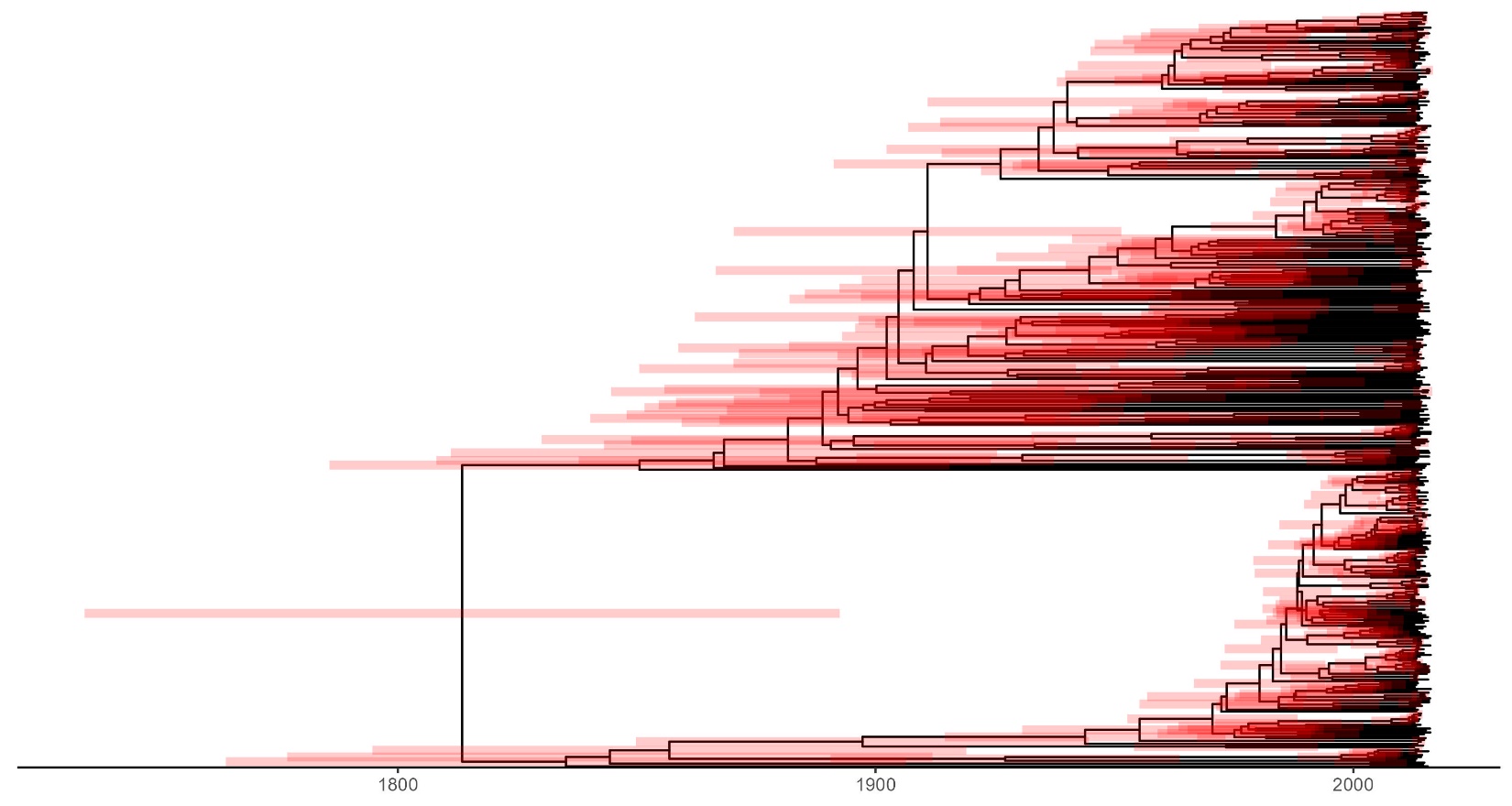

Supplementary Figure 2g: Maximum clade credibility tree of *Mycobacterium tuberculosis* complex lineage 4.4 with 95% highest posterior density intervals of node heights

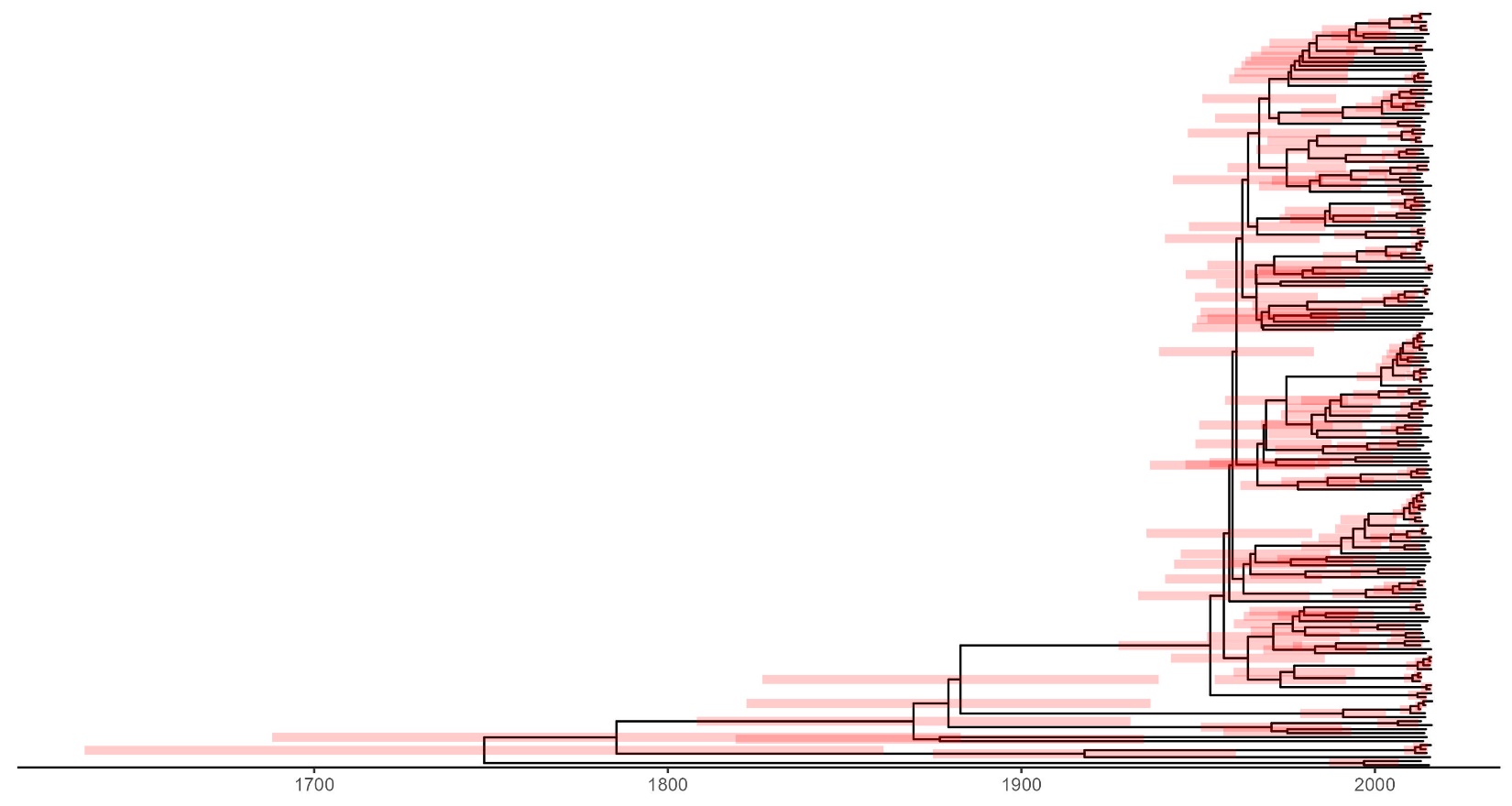

Supplementary Figure 2h: Maximum clade credibility tree of *Mycobacterium tuberculosis* complex lineage 4.8 with 95% highest posterior density intervals of node heights

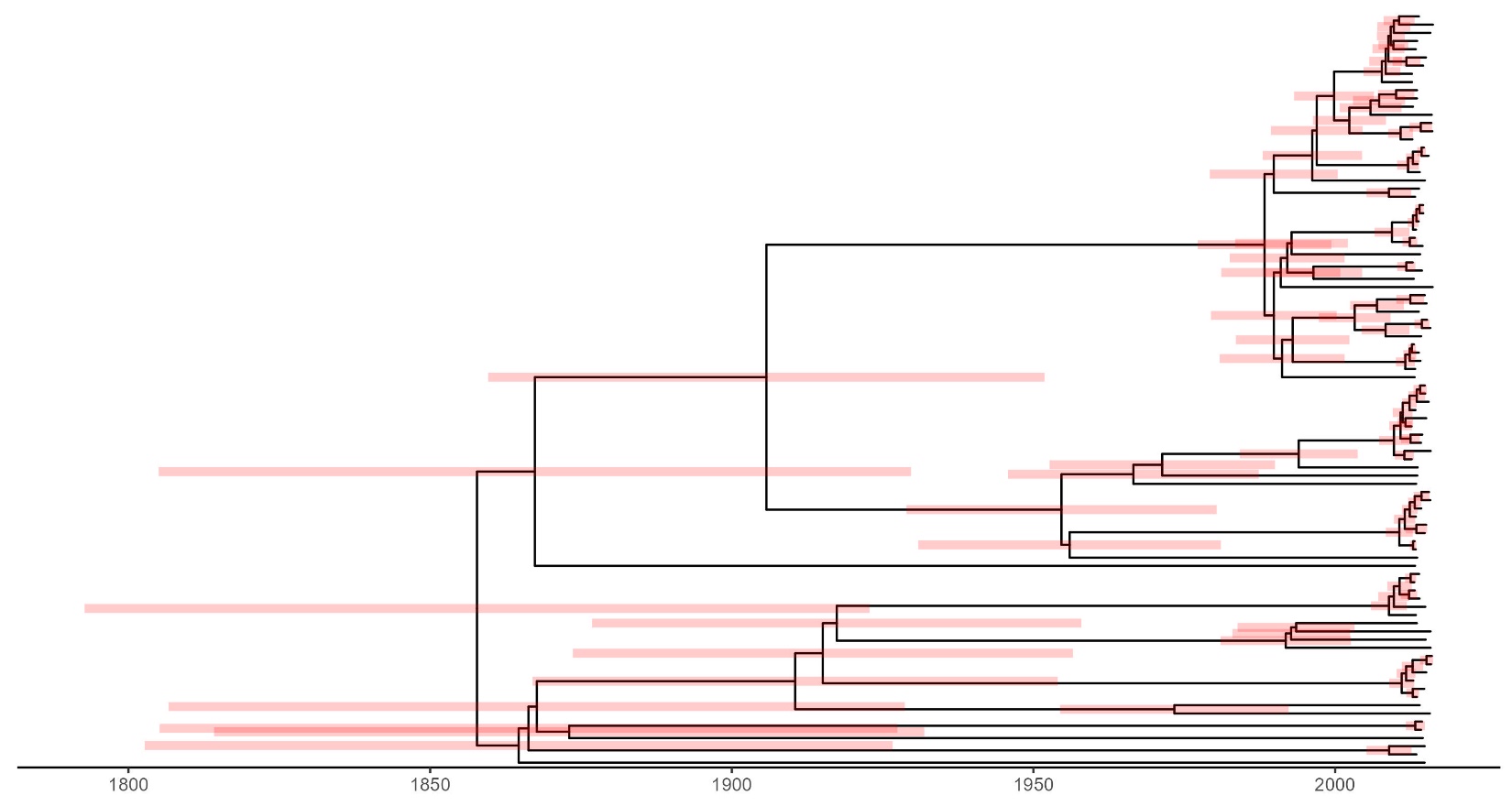

Supplementary Table 1. Posterior clock rate of the *Mycobacterium tuberculosis* complex lineages

| **Lineage** | **Clock rate (s/s/y)**  **posterior mean** | **Clock rate (s/s/y)**  **posterior median** | **95% HPD interval** |
| --- | --- | --- | --- |
| 1 | 3.4E-7 | 3.3E-7 | 2.1E-7, 4.6E-7 |
| 2 | 3.7E-7 | 3.6E-7 | 2.4E-7, 5.0E-7 |
| 4.1.1 | 1.4E-7 | 1.4E-7 | 8.0E-8, 2.0E-7 |
| 4.1.2 | 1.3E-7 | 1.3E-7 | 7.8E-8, 1.9E-7 |
| 4.3.2 | 2.0E-7 | 2.0E-7 | 1.3E-7, 2.7E-7 |
| 4.3.4 | 1.0E-7 | 1.0E-7 | 6.5E-8, 1.4E-7 |
| 4.4 | 1.5E-7 | 1.5E-7 | 9.2E-8, 2.1E-7 |
| 4.8 | 2.0E-7 | 2.0E-7 | 1.3E-7, 2.9E-7 |

Abbreviations: s/s/y: substitutions per site per year. HPD: highest posterior density.

Supplementary Table 2. Genomic cluster proportions (based on a 12-SNPs cutoff) of the *Mycobacterium tuberculosis* complex lineages

| **Lineage** | **Cluster proportions** | **Crude OR (95% CI)** | **Adjusted^1^ OR (95% CI)** |
| --- | --- | --- | --- |
| L1 | 38/86 | 1 | 1 |
| L2 | 57/72 | 4.80 (2.40, 10.01) | 4.44 (2.19, 9.37) |
| L4.1.1 | 103/141 | 3.42 (1.96, 6.07) | 3.48 (1.97, 6.25) |
| L4.1.2 | 96/149 | 2.29 (1.34, 3.95) | 2.08 (1.20, 3.64) |
| L4.3.2 | 79/117 | 2.63 (1.48, 4.70) | 2.52 (1.41, 4.55) |
| L4.3.4 | 260/400 | 2.35 (1.47, 3.78) | 1.49 (0.91, 2.47) |
| L4.4 | 124/189 | 2.41 (1.44, 4.08) | 2.21 (1.30, 3.79) |
| L4.8 | 73/92 | 4.85 (2.54, 9.57) | 4.32 (2.24, 8.62) |

Abbreviations: SNP: single nucleotide polymorphism. OR: odds ratio. CI: confidence interval.

^1^Adjusted for age, gender, HIV status, and district.
